# Cataract Surgery Outcomes and Postoperative Patient Compliance in Limited English Proficiency Patients at a County Hospital

**DOI:** 10.1101/2021.12.01.21267163

**Authors:** Colleen C. Yard, Kayla R. Walter, Ning O. Zhao, Alice Z. Chuang, Kimberly A. Mankiewicz, Eric L. Crowell

**Affiliations:** Ruiz Department of Ophthalmology and Visual Science, McGovern Medical School at The University of Texas Health Science Center at Houston (UTHealth), Houston, TX, USA; Robert Cizik Eye Clinic, Houston, TX, USA; Lyndon B. Johnson Hospital, Harris Health, Houston, TX, USA

**Author notes:** **Corresponding Author:** Eric L. Crowell, MD, MPH, 1601 Trinity St., Bldg. B Z1200 Suite 3.706, Austin, TX, 78712. **Disclosures:** None.

**Keywords:** Limited English proficiency, language, cataract, county hospital

## Abstract

**Background/Aims:** Investigate the role of language barriers in cataract surgery outcomes at a county hospital.

**Methods:** Retrospective chart review of patients who underwent cataract surgery March 2018-February 2019 at Lyndon B. Johnson Hospital. Patients who underwent cataract surgery combined with another procedure or had severe glaucoma or proliferative diabetic retinopathy were excluded. Patients were classified into limited English proficient (LEP) or English proficient (non-LEP) groups based on language preferences. Demographics, baseline ocular characteristics, intraoperative complications, postoperative BCVA (best-corrected visual acuity), complications, and compliance were recorded. The primary outcome was incidence of poor visual outcomes (BCVA<20/40) at the postoperative 1-month visit.

**Results:** 354 patients (199 [56%] LEP and 155 [44%] non-LEP) with 125 (35%) males and a mean age 66.1 (±10.9) years were included. LEP patients were about 5 years older than non-LEP patients (*P*<0.001) and were mostly Hispanic (172 [86%] LEP vs. 36 [26%] non-LEP, *P*<0.001). The baseline ocular characteristics were similar (*P* >0.05), except severity of cataract (125 [63%] NSC grade >2+ for LEP vs 70 [51%] for non-LEP, *P* =0.03). No significant differences in intraoperative complications (*P* =0.18), incidence of poor vision (*P* =0.59), postoperative cystoid macular edema (*P* =0.32), and compliance with the postoperative drop regimen (*P* =0.11) were noted.

**Conclusion:** There were no statistically significant differences in incidence of poor vision, complications, or compliance. However, there was a trend toward significance, showing that language barriers may lead to more advanced disease and compliance issues with postoperative medications.

**SYNOPSIS:** Language barriers may lead to compliance issues with postoperative medication regimens, as shown by the difference in postoperative care adherence rates between limited English proficient and English proficient patients.

## INTRODUCTION

With over 24.4 million Americans affected, cataract is the leading cause of blindness worldwide in adults 50 years and older [1]. Phacoemulsification cataract surgery with intraocular lens implantation is routinely performed in the United States to restore vision. However, as with any surgery, there are complications, which include retinal detachment (0.8%) [2] and endophthalmitis (0.1%) [3]. Risk factors that affect postoperative outcomes include poor preoperative visual acuity, ocular history, age, sex, and race [4-9].

In preparation for surgery, there is an extensive preoperative evaluation process, which is frequently complicated by language and cultural differences. Harris Health System, the county health system for Houston and surrounding Harris County, serves a population where 50% of patients are of Limited English Proficiency (LEP). LEP is defined as the inability to speak, read, write, and/or understand the English language at a level that prevents the patient from interacting effectively with health care providers [10].

The effect of LEP on successful medical care has been previously investigated in other specialties. A systematic review of studies across many disciplines found that the use of an interpreter, both *ad hoc* and licensed medical interpreters, resulted in improved patient outcomes as well as patient satisfaction [11]. Furthermore, Parker et al showed that Spanish-speaking patients who had a physician fluent in Spanish had a 10% higher rate of adherence to their diabetic medications than the Spanish-speaking patients who had an English-speaking physician [12]. LEP is an obvious barrier to effective clinical care, and an investigation on the effects in all specialties, including Ophthalmology, is warranted.

Understanding the effect of LEP is even more crucial when the decision is made to proceed with surgery. During the pre-, intra-, and postoperative periods, understanding and cooperation with instructions can affect the patient’s outcome. Although this has not been studied in Ophthalmology, a study from the Ear, Nose, and Throat (ENT) specialty compared the complication and compliance rates with postoperative instructions, written in English or Spanish, in an LEP pediatric ENT population. After surgery, the patients’ families were randomized to receive written postoperative instructions in English or Spanish. Two patients who received the English instructions noted minor postoperative complications. The study was only conducted on 11 patients, but even in the small population, it is clear that the families preferred instructions in their native language. Most notably, the patients who received written Spanish instructions were more satisfied with the overall experience [13]. This study provides insight into how the language barrier issues in the clinic translate similarly to the perioperative environment.

Language barrier issues have never been investigated in the field of Ophthalmology. Our study compared the baseline ocular characteristics, rates of intra- and postoperative complications, patient compliance rates, and visual outcomes between English-speaking and LEP patients who underwent cataract surgery at our county health system. We chose cataract surgery to investigate due to its prevalence and the extensive communication that occurs during the pre-and postoperative visits. The goal of this research is to promote better access to language tools in the clinic and perioperative environment.

## METHODS

This retrospective chart review was conducted at the Lyndon B. Johnson General Hospital (LBJ) of the Harris Health System in Houston, TX, USA. Institutional Review Board (IRB) approval was obtained from The University of Texas Health Science Center Committee for the Protection of Human Subjects and the Harris Health System. All research adhered to the tenets of the Declaration of Helsinki and was HIPAA compliant. The IRB determined informed consent could be waived.

### Study Population

Charts of patients who underwent cataract surgery between March 2018 and February 2019 at LBJ and had follow-up for at least 1 month were reviewed. Eyes that underwent combined procedures or those with proliferative diabetic retinopathy, severe glaucoma, or ocular trauma were excluded. If both eyes met the criteria, the first eye was included. Patients were divided into LEP and non-LEP groups based on the documented native language.

### Data Collection

Data collected included demographics (age, sex/ethnicity, race), baseline ocular characteristics (nonproliferative diabetic retinopathy [NPDR], mild or moderate glaucoma, type and grade of cataract), whether a complex vs. simple cataract surgery was performed (as defined by the CPT codes 66982 and 66984, respectively), and intraoperative complications (including broken capsular bag, retained lens fragments, etc.). Postoperative data were collected at Day 1 and Weeks 1, 4, and 8, including best-corrected visual acuity (BCVA), drop compliance, and postoperative complications (including pseudophakic cystoid macular edema, retinal tear or detachment, endophthalmitis, etc.).

### Data Analysis

Poor visual outcome was defined as Snellen BCVA worse than 20/40 at the last study visit. Data were summarized by mean (± standard deviation) for continuous variables or by frequency (%) for discrete variables. Two-sample *t*-test and Fisher Exact tests were used to compare variables between LEP and non-LEP groups. All statistical analyses were performed using SAS for Windows 9.4 (SAS Inc, Cary, NC). A *P* value less than 0.05 was considered statistically significant.

## RESULTS

As seen in Table 1, 354 (199 [56%] LEP and 155 [44%] non-LEP) patients with a mean age 66.1 (±10.9, range 31 - 95) years were included. One hundred and twenty-five (125; 35%) were males. All LEP and 140 non-LEP patients reported race/ethnicity. In the LEP patients, 172 (86%) were Hispanic, 21 (11%) Asian, 3 (2%) White, and 3 (2%) Black. For non-LEP patients, 36 (26%) were Hispanic, 9 (6%) Asian, 65 (46%) White, and 33 (10%) Black (*P*<0.001). There was no difference in the sex distribution between groups; however, age and race/ethnicity distributions were significantly different between groups (Table 1). LEP patients were about 5 years older than non-LEP patients (*P*<0.001) and were mostly Hispanic (86%).

**Table 1.** Demographics and Baseline Ocular Characteristics

| Variable | All<br>(N=354) | Non-LEP<br>(N=155) | LEP<br>(N=199) | P |
| --- | --- | --- | --- | --- |
| **Demographics and Baseline Characteristics** |  |  |  |  |
| Age (years, SD [min, max]) | 66.1 (10.9)<br>[31 – 95] | 62.9 (9.9)<br>[31 – 95] | 68.5 (11.1)<br>[32 – 92] | <0.001 |
| Sex (males, %) | 125 (35%) | 55 (35%) | 70 (35%) | 1.0 |
| Race/ethnicity (no. reported, %) | 339 (96%) | 140 (90%) | 199 (100%) |  |
| White (%) | 68 (20%) | 65 (46%) | 3 (2%) | <0.001 |
| Black (%) | 33 (10%) | 30 (21%) | 3 (2%) |  |
| Hispanic (%) | 208 (61%) | 36 (26%) | 172 (86%) |  |
| Asian (%) | 30 (9%) | 9 (6%) | 21 (11%) |  |
| Second eye underwent cataract surgery (%) | 83 (23%) | 35 (23%) | 48 (24%) | 0.80 |
| Complicated cataract surgery in the first eye (%) | 22 (27%) | 10 (29%) | 12 (25%) | 0.80 |
| Nonproliferative diabetic retinopathy (%)<sup>1</sup> | 55 (16%) | 23 (15%) | 32 (16%) | 0.88 |
| Glaucoma (non-severe, %) | 18 (5%) | 4 (3%) | 14 (7%) | 0.086 |
| *Cataract Type and Grade* |  |  |  |  |
| Nuclear (%) | 329 (93%) | 142 (92%) | 187 (94%) | 0.41 |
| Grade > 2+ (%) | 204 (58%) | 79 (51%) | 125 (63%) | 0.030 |
| Cortical (%) | 191 (54%) | 90 (58%) | 101 (51%) | 0.20 |
| Grade > 2+ (%) | 73 (21%) | 42 (27%) | 31 (16%) | 0.012 |
| Posterior capsular (%) | 187 (53%) | 81 (52%) | 106 (53%) | 0.91 |
| Grade > 2+ (%) | 84 (24%) | 36 (23%) | 48 (24%) | 0.90 |
| Underwent baseline OCT exam (%) | 149 (42%) | 62 (40%) | 87 (44%) |  |
| No View (%) | 29 (20%) | 13 (21%) | 16 (19%) | 0.92 |
| Macular edema on OCT exam (%) | 24 (16%) | 10 (16%) | 14 (16%) |  |

<sup>1</sup> missing 8 data points, 4 in English group, 4 in non-English group SD=standard deviation; LEP=limited English proficiency

### Baseline Ocular Characteristics

As seen in Table 1, 271 eyes (77%) were the first eye of each patient to undergo cataract surgery. Of the 83 eyes that were “second eyes,” 22 (27%) had complicated cataract surgery in the previous eye. Fifty-five (55; 16%) of 346 eyes had NPDR. Rates of complex first eye surgery and NPDR were not different between the LEP and non-LEP groups (*P*=0.80 and *P*=0.88, respectively). There were 18 (5%) patients with mild or moderate glaucoma, 14 (7%) in LEP and 4 (3%) in non-LEP groups. Although the rate of non-severe glaucoma was not significantly different between the LEP and non-LEP groups (*P*=0.086), the LEP group had 4% more patients with glaucoma when compared to the non-LEP group.

Three hundred and twenty-nine (329; 93%) eyes had nuclear sclerotic cataracts (NSC), 191 (54%) had cortical cataracts (CC), and 187 (53%) had posterior subcapsular cataracts (PSC). The types of cataracts were not significantly different between groups (*P*>0.20). The percent of eyes with NSC grade > 2+ was significantly higher in the LEP group (125 [63%] for LEP vs. 70 [51%] for non-LEP, *P*=0.03), while the percent of eyes with CC grade > 2+ was significantly less in LEP (31 [16%] for LEP vs. 42 [27%] for non-LEP, *P*=0.012). Of the 149 eyes (42%) that underwent ocular coherence tomography (OCT) examination (87 [44%] for LEP and 62 [40%] for non-LEP), the rates of macular edema were the same (16%, 14 LEP patients and 10 non-LEP patients) between groups (Table 1).

### Cataract Surgery

As seen in Table 2, only 26 (13%) patients in the LEP group had any documented use of an interpreter at one of the at least 5 pre- and postoperative visits. This refers to all the visits related to the preoperative process. This percentage of interpreter usage is likely lower than reality because this data was mined from the electronic medical records (EMR). The EMR only automatically records phone interpreter encounters. Our clinic physicians often use an in-person interpreter assigned to our clinic, and that is not recorded in the EMR. Furthermore, many of our physicians are bilingual and do not need an interpreter for the basic clinical exam. In our health system, an interpreter is required for the consent process, but with the extensive use of in-person interpreters, that is not always recorded except on the consent form. Complex cataract surgery occurred in 100 (50%) LEP and 71 (46%) non-LEP (*P*=0.45) patients, with 9 (5%) LEP and 13 (8%) non-LEP patients experiencing intraoperative adverse events (*P*=0.18). A dense cataractous lens was the most frequently noted explanation for complex cataract surgery (133 [38%]). Although it was not statistically significant between groups (*P*=0.077), the LEP group had a 10% greater prevalence of dense cataracts when compared to the non-LEP group (83 [42%] for LEP and 50 [32%] for non-LEP). Usage rates of complex surgery tools were not different between groups. Specifically, iris-expansion devices were used in 26 eyes (7%), capsular tension rings in 5 eyes, (1%), and Trypan blue in 143 eyes (40%).

**Table 2.** Summary of Parameters during Cataract Surgery

| **Variable** | **All**<br>**(N=354)** | **Non-LEP**<br>**(N=155)** | **LEP**<br>**(N=199)** | **P** |
| --- | --- | --- | --- | --- |
| Interpreter Used | 26 (7%) | 0 (0%) | 26 (13%) | <0.001 |
| Complex surgery (%) | 171 (48%) | 71 (46%) | 100 (50%) | 0.45 |
| Dense lens | 133 (38%) | 50 (32%) | 83 (42%) | 0.077 |
| Small pupil | 19 (5%) | 9 (6%) | 10 (5%) | 0.81 |
| Floppy iris | 8 (2%) | 2 (1%) | 6 (3%) | 0.47 |
| Anterior capsular tear | 7 (2%) | 4 (3%) | 3 (2%) | 0.70 |
| Zonular loose/weakness/dehiscence | 10 (3%) | 4 (3%) | 6 (3%) | 1.00 |
| Polar cataract | 3 (1%) | 2 (1%) | 1 (1%) | 0.58 |
| Uveitis | 3 (1%) | 2 (1%) | 1 (1%) | 0.58 |
| Use of iris manipulation device (%) | 26 (7%) | 14 (9%) | 12 (6%) | 0.31 |
| Use of capsular tension ring (%) | 5 (1%) | 2 (1%) | 3 (2%) | 1.00 |
| Use of Trypan blue (%) | 143 (40%) | 59 (38%) | 84 (42%) | 0.45 |
| Use of other tools | 8 (2%) | 5 (3%) | 3 (2%) | 0.31 |
| Intraoperative complications | 22 (6%) | 13 (8%) | 9 (5%) | 0.18 |
| Capsular bag rupture (%) | 22 (6%) | 13 (8%) | 9 (5%) | 0.18 |
| Retained lens material (%) | 6 (2%) | 3 (2%) | 3 (2%) | 1.0 |

LEP=limited English proficiency

Intraoperative complications occurred in 22 eyes (6%), and there was no difference between groups (9 [5%] for LEP and 13 [8%] for non-LEP, *P*=0.18). All complications were due to capsular bag rupture.

### Outcomes

As demonstrated in Table 3, poor visual outcomes occurred in 36 (18%) LEP and 32 (21%) non-LEP patients (*P*=0.59). Postoperative cystoid macular edema occurred in 45 (23%) LEP and 43 (28%) non-LEP patients (*P*=0.32). No endophthalmitis, retinal tears, or detachments occurred. Compliance with the postoperative drop regimen occurred at a rate of 81% (161 eyes) in the LEP group and 88% (136 eyes) in the non-LEP group (*P*=0.11). Although not statistically significant, it was one of our secondary outcome measures and did trend towards significance.

**Table 3.** Outcomes

| **Variable** | **All**<br>**(N=354)** | **Non-LEP**<br>**(N=155)** | **LEP**<br>**(N=199)** | ***P*** |
| --- | --- | --- | --- | --- |
| Poor vision (> 20/40, %) | 68 (19%) | 32 (21%) | 36 (18%) | 0.59 |
| Post cataract CME (%) | 88 (25%) | 43 (28%) | 45 (23%) | 0.32 |
| Drop compliance (No, %) | 297 (84%) | 136 (88%) | 161 (81%) | 0.11 |

LEP=limited English proficiency; CME=cystoid macular edema

## DISCUSSION

This study sought to address a fundamental problem in many of our academic medical centers in this age of globalization: the challenge of communicating with our diverse patient population. This is of particular importance to ophthalmologists, as eye surgery is mostly performed under light sedation, where communication is essential for good outcomes. Although cataract surgery is one of the most widely performed surgeries in the world, with over 10 million surgeries performed each year [14], most cataract surgery outcomes studies focus on different surgical techniques, surgeon volume, different postoperative management strategies, or rates of complications [2, 5, 7, 8] and do not consider the reasons behind these complications. This is the first study in Ophthalmology to provide surgical outcomes analysis that takes into account the effect of a language barrier (PubMed search on August, 12 2020, using a combination of terms cataract, surgery, language, limited, English, proficiency). The only other similar report that we found was an anecdotal article. In this article, the author highlights how LEP is clearly an issue with Ophthalmology patients by describing a patient interaction where a language barrier impeded appropriate postoperative care [15].

Our LEP patient population was notably older and consistently had denser cataracts documented preoperatively (Table 2). More LEP patients also had glaucoma, making them possibly more susceptible to complications in cataract surgery (Table 2). These 2 characteristics demonstrate that LEP patients may be more hesitant to visit an ophthalmologist, and thus wait to address the issue until the cataract is very advanced and detrimental to their daily life. This idea has previously been explored in Internal Medicine. Alzubaidi et al demonstrated that LEP Arabic-speaking patients were less likely to seek care than their English-speaking counterparts, which can lead to more severe ailments and complications [16]. This concept is important when considering the burden that poor, correctable, vision places on society.

The incidence of complex surgery was high in both groups in our study, about 50%. For comparison, in a 2019 population-based study of Medicare patients, 95% of cataract surgeons performed complex surgery less than 35% of the time [17]. The complexity of our study’s surgeries was disproportionally high due to dense lenses (42% for LEP versus 32 % for non-LEP, Table 2). This lens density is likely because of the older age of the patients and, therefore, more advanced disease.

The use of the other complex surgery devices, such as iris-expansion devices and capsular tension rings, was the same between the groups; however, there are many intangibles during surgery that are not documented in an operative report, such as the need to constantly redirect a patient who does not understand instructions during the surgery. These “intangibles” can make surgery more challenging but are seldom mentioned in an operative report.

Additionally, these challenges can add significant time to surgery, which consequently increases anesthesia risk and potential surgical costs. However, these aspects of the surgical process were outside of the scope of this study.

There was no statistically significant difference between the LEP and non-LEP groups in visual outcomes, rates of complications, or drop compliance. Although there was not a statistically significant difference, drop compliance was 7% higher in the non-LEP group; however, our study was not powered to detect a difference of less than 10%. Of all the intraoperative and postoperative metrics that were collected, compliance is the metric from this study most dependent on verbal communication. While the metric most dependent on communication did not reach statistical significance between our groups, it was the only metric that trended towards significance. This implies that poor communication may indeed influence drop compliance in a higher-powered study.

## Limitations

Our retrospective study relied on the review of an operative report to detail the operative picture, and patient compliance with the medication regimens was self-reported.

Another major weakness is the heterogeneity of the terms “LEP” and “non-LEP”. This was defined by using the primary language that was documented in the patient’s electronic medical record. However, a patient’s primary language may have been listed in the electronic medical record as English, when, in reality, he or she was actually a very limited English speaker. These nuances were impossible to elicit with a chart review. Anecdotally, most patients who had Spanish listed in the EMR were not proficient in English based on our experiences in the clinic.

Our study is not the first study to have difficulty defining LEP in the clinical setting. There is no standardized questionnaire in the medical literature. If such a questionnaire were to exist, it would help provide a more qualitative and consistent way to define a patient as LEP. The only standard that has been used before has been the United States Census questionnaire. In one study, they copied the census question, which asked, “Do you speak English? (1) Very well (2) Well (3) Not well (4) Not at all.” They found that any patient that answered anything less than “very well” usually needed an interpreter. The way they defined this need was if the patient needed significant help in discussing their care with the physician. Difficulty having this type of discussion correlated very highly with answering anything except “very well” in the survey [18]. This survey is something we could consider employing in a future prospective study.

A prospective study would improve some of the other study limitations as well. For one, the surgery could be observed, allowing for better intraoperative reports. Prospective data collection would ensure that a patient was truly LEP, and designated interpreters could be used throughout the study to eliminate communication inconsistencies.

## Conclusion

Overall, there was no statistically significant difference in any of the major outcomes between the 2 groups. However, there is a trend toward significance, showing that language barriers may lead to more advanced disease and compliance issues with postoperative medication regimens. Although none of the outcome metrics showed statistical significance, our study highlights the importance of physician communication and the need for clear, interpreter-mediated conversations. In our ever-diversifying country and world, this need has become ever present, which our research underscores. However, a higher-powered, prospective study is needed to investigate this relationship further.

## Data Availability

All data produced in the present study are available upon reasonable request to the authors.

